# Effectiveness of integrated care for older adults with depression and hypertension in rural China: a cluster randomized controlled trial

**DOI:** 10.1101/2022.05.16.22275122

**Authors:** Shulin Chen, Yeates Conwell, Jiang Xue, Lydia Li, Tingjie Zhao, Wan Tang, Hillary Bogner, Hengjin Dong

## Abstract

**Background:** Effectiveness of integrated care management for common, comorbid physical and mental disorders has been insufficiently examined in low- and middle-income countries. We tested hypotheses that older adults treated in rural Chinese primary care clinics with integrated care management of comorbid depression and HTN would show greater improvements in depression symptom severity and hypertension (HTN) control than those who received usual care.

**Methods and findings:** The study was a 12-month cluster randomized controlled trial conducted from 2014 through 2017, with analyses conducted in 2020-2021. Subjects were rural village clinics of randomly selected towns in Zhejiang Province, China. Ten towns with a total of 218 rural village primary care clinics were randomized, five towns each, to deliver the Chinese Older Adult Collaborations in Health (COACH) intervention or enhanced care-as-usual (eCAU). The COACH intervention consisted of algorithm-driven treatment of depression and HTN by village primary care doctors supported by village lay workers with consultation from centrally-located psychiatrists. Subjects included clinic patients aged ≥60 years with a diagnosis of HTN and clinically significant depressive symptoms (PHQ-9 score ≥10). Of 2899 eligible subjects, 2365 (82%) agreed to participate and were followed for 12 months. Observers were blinded to study hypotheses but not to group assignment. Primary outcomes specified a priori were change in depression symptom severity and proportion with controlled HTN.

Compared with 1133 subjects who received eCAU, 1232 COACH subjects showed greater reduction in depressive symptoms (Cohen’s d [±SD] = -0.21 [-0.25, -0.17]) and greater likelihood of achieving HTN control (OR [95% CI] = 18.24 [8.40, 39.63]). Exploratory post hoc analyses showed that COACH subjects who accepted an antidepressant had greater symptom reduction than either those who declined the medication or received eCAU. HTN control improved in COACH subjects regardless of antidepressant use.

**Conclusions:** The COACH model appears effective in managing comorbid depression and HTN in older adult residents of rural Chinese villages. Integrated care management of comorbid depression and common medical illness may be a useful approach in other low resourced settings in which specialty geriatric mental health care is lacking.

## Introduction

Older adults are the fastest growing segment of the world’s population, a pattern especially pronounced in low and middle-income countries (LMICs). China has an estimated 254 million people over the age of 60 years [1]. Depression and hypertension (HTN) are among the most common disorders of later life [2,3]. Known as “the silent killer” [4], HTN is a major cause of strokes and ischemic heart disease, while depressive disorders are the third leading cause of years lived with disability worldwide [5]. Depression and HTN commonly coexist, making their management more complex and clinical outcomes worse [6-11].

Primary care providers (PCPs) in rural China receive little training in the detection, diagnosis, and management of common mental disorders and mental health specialty care is difficult to access [6,12]. Consequently, late life affective disorders are rarely treated. In a naturalistic longitudinal study of older urban residents in China, Chen and colleagues found that only 1% of those with a major depressive episode received treatment over 12 months [13].

In Western countries collaborative depression care management models [14-16] have proven effective in managing depression and comorbid medical conditions while also reducing costs in primary care settings [17-19]. Rarely, however, have studies been conducted in LMICs that used a randomized control trial (RCT) design, had a focus on older adults, or examined comorbid depression and medical illness [20-22].

We compared a collaborative depression care management intervention called Chinese Older Adult Collaborations in Health [COACH] to enhanced care as usual (eCAU) for the treatment of comorbid depression and HTN in older residents of rural Chinese village clinics. Our primary hypotheses were that older adults with comorbid depression and HTN who received the COACH intervention would show greater improvements in both depressive symptom severity and HTN control than those who received eCAU over 12 months. We also explored *post hoc* whether acceptance of antidepressant medications by those in the COACH intervention was associated with treatment response.

## Methods

### Study design

The COACH Study is a cluster RCT conducted in Tonglu and Jiande counties of Zhejiang Province, China. Together these two counties have a total of 920,000 residents distributed across 30 towns with a total of 602 villages. The health needs of each village are provided by a clinic staffed by one PCP without other nursing support. Mental health care is provided by one county-level mental hospital and residents’ social needs are provided by the village’s Aging Association staffed by local residents.

The study was reviewed and approved by the institutional review boards of Zhejiang University, the University of Rochester, and the U.S. National Institute of Mental Health (NIMH). Further details on the trial design, procedures, and statistical analysis plan are available elsewhere [23].

### Randomization and masking

The original design planned for village to be the unit of randomization [23]. However, it became clear prior to initiation of the study that the chance of contamination bias between study conditions was substantial due to interactions between neighboring village PCPs. Therefore, randomization was based on the town while outcomes pertain to the individual level. Ten towns in Tonglu and Jiande County, each containing from 18 to 25 villages, were randomly selected by a computer algorithm administered by the study statistician to assure that no two shared a common boundary. Five towns were selected for each arm of the study, with all villages and their associated clinics in each town being assigned either to deliver the COACH intervention or eCAU to eligible patients. The PCP and aging workers (AW) for COACH intervention clinics (see “Intervention-COACH” below) were approached by the research team for agreement to participate. One village assigned to the COACH arm refused. The final numbers of villages in the COACH and eCAU intervention arms were 102 and 116 respectively.

Masking was not feasible for research participants or assessors. All were blinded, however, to the study objectives and hypotheses.

### Participants

Inclusion criteria for subjects were registration in the village’s primary care clinic; age ≥60 years; clinically significant depression defined as a Patient Health Questionnaire-9 (PHQ-9) total score of ≥10 [24]; chart diagnosis of HTN; intact cognitive functioning (Six-Item Screener [SIS] score <3) [25]; and willing to give written informed consent. Exclusion criteria included mania, psychosis, or alcohol abuse or dependence active in past 6 months based on the MINI Diagnostic Interview [26]; and acute suicide risk. Subjects in the COACH group could decline antidepressant prescription and remain in the study. Potentially eligible subjects were invited to meet in their homes or the clinic with a research assistant (RA) who introduced the study, assessed eligibility, and obtained written informed consent.

### Assessment

Trained research assessors administered study measures in the subject’s home or the clinic at study entry and 1, 3, 6, 9, and 12 months later. In addition to sociodemographic information, measures assessed quality of life using the WHOQOL-BREF [27,28], the number of medical comorbidities recorded in the subject’s medical record, impairment in basic (ADL) and instrumental activities of daily living (IADLs) [29], social support using the Chinese version of the Medical Outcomes Study Social Support Survey (MOS-SSS-C) [30] and social network size [31].

The primary measure of depressive symptoms was total score on a valid and reliable Chinese translation of the 17-item Hamilton Depression Rating Scale (HDRS) [32]. BP was measured at baseline and each follow-up point according to China CDC standards [23]. The primary HTN outcome was uncontrolled HTN, defined as systolic BP ≥130 mmHg or diastolic BP ≥80 for patients with diabetes mellitus, coronary heart disease or renal disease, and systolic BP ≥140 or diastolic BP ≥90 for all others [33].

### Comparison condition -- Enhanced Care-as-Usual (eCAU)

PCPs in village clinics ordinarily have three years of medical education after high school [34]. They receive periodic in-service education that includes HTN management using CDC practice guidelines [35]. When PCPs suspect mental illness in their patients, they ordinarily recommend the patient go to the County Mental Hospital. PCPs cannot initiate antidepressant treatment, but can renew prescriptions initiated by psychiatrists.

We refer to care as CAU as “enhanced” (eCAU) because PCPs were told when their patients screened positive for depression and were provided with copies of antidepressant treatment guidelines adapted from the Duke Somatic Treatment Algorithm for Geriatric Depression (STAGED) [36].

### Intervention – COACH

Training for the COACH team in each village, which consisted of the clinic’s PCP, the village AW, and a county hospital Psychiatrist, took place over four days prior to the intervention. It included instruction in their respective roles and how to work collaboratively. PCPs received training in depression assessment, case management using the toolkit adapted from the MacArthur Initiative on Depression in Primary Care [37], and use of the STAGED antidepressant treatment guidelines [22,24,36]. There were no restrictions on concomitant care or treatment.

The AWs’ role was to educate subjects about their conditions followed by goal setting and collaborative development of behavior change strategies. Their training curriculum included an overview of depression, HTN, and their relationship; principles of disease self-management; psychosocial assessment and care planning; psychoeducation with older adults and their families; and ethical standards including confidentiality. They did not receive training in manualized psychosocial interventions. During home visits, AWs helped the older person to set behavioral goals, review progress, and modify the plan if needed. In addition, the AWs created social opportunities for older persons, for example, by organizing social activities and educational workshops.

### Intervention procedures and outcomes

The psychiatrist traveled to each village clinic where they conducted assessments of eligible subjects’ depression status in consultation with the PCP. The AW assessed the person’s social supports, lifestyle, and functional, nutritional, and financial status. The AW, PCP and psychiatrist reviewed all the assessment data and constructed a care plan.

The PCP and AW met with each subject monthly to monitor BP and depressive symptoms, assess antidepressant and antihypertensive use and side-effects, and provide support to the patient and family. The PCP and AW met weekly in person to review their shared caseload, and with the Psychiatrist monthly by telephone to assess progress and update the treatment plan as indicated.

Primary outcomes of the study were change in depression symptom severity as measured by the HDRS at baseline and follow up assessments, and the proportions (%) of subjects whose HTN was controlled. Initial plans to measure adherence to medications could not be completed for administrative reasons.

### Statistical analysis

The study aimed to recruit 1200 subjects to each study arm. Because the intra-class correlation (ICC) at the town level was likely to be low, the study used a 3-level nested design in which the power was based on the ICC among the patients within the village and serial correlation between repeated assessments within the patient. With this number of subjects, setting the serial correlation at 0.5 and varying the ICC over 0.05, 0.1 and 0.2, and assuming a two-sided type I error = 0.05, power = 0.8 and an attrition rate of 20%, the detectable effect size ranged from 0.17 to 0.26 for the continuous outcome of depression symptom change. For the dichotomous outcome of HTN control, we also set the serial correlation at 0.5 and varied the percent of variance between patients within the village clinic over 0.05, 0.1 and 0.2. Based on two-sided type I error = 0.05, power = 0.8, base rate 0.5 (most conservative) and 20% attrition rate, the detectable between-group proportion with 2400 subjects ranged from 11% to 17%, well within the range of clinically meaningful differences in primary care settings.

We compared baseline characteristics between groups using t-test and chi-square. Characteristics significantly differentiating the two groups at p<.05 were then treated as covariates when testing between-group differences using longitudinal models.

To test our hypotheses that COACH participants would show greater improvements in depressive disorders and HTN control than eCAU participants over 12 months of involvement in the study, we modeled the repeatedly assessed variables of depressive symptom severity and of HTN control using generalized linear mixed effect models (GLMM) and weighted generalized estimating equations (WGEE). Since the GLMM and WGEE were consistent, only the GLMM results were reported [38]. Analyses were performed as intention-to-treat using all subjects recruited from the ten towns being randomized. Random effects of towns, villages, and subjects were used to account for the within-cluster associations within the towns and villages, as well as repeated measurements within the subjects. For each outcome, time and intervention were predictors adjusting for covariates. We assessed the potential interaction between time and intervention using linear contrasts to assess COACH vs. eCAU differences over the 12-month period as well as any sub-intervals within this period.

With regard to model assumptions, the functional forms or scales of the quantitative covariates were examined using the fractional polynomial approach of Royston and Altman [39], and the final models were selected based on the Akaike information criterion. We further calculated the R squared, both marginal and conditional, as proposed by Vonesh et al [40] to assess the goodness-of-fit of these selected generalized linear mixed effect models. The marginal R squared ranged from 0.24 to 0.29 and the conditional R squared ranged from 0.65 to 0.83, all of which fall in the acceptable range for psychosocial studies. In order to check the normality of residuals we used QQ-plot to demonstrate that all models were normally distributed with constant variance.

Following completion of analyses planned *a priori*, we conducted a *post hoc* descriptive comparison of subjects within the COACH group who had accepted prescription of an antidepressant as a component of their treatment (Antidep[+]) with those who had not (Antidep[-]). The purpose was to explore whether antidepressant use may explain observed differences in depression and HTN treatment response between the two subgroups.

Statistical analyses were conducted with Statistical Analysis System software version 9.4 or above. The study protocol was prospectively registered with Grants.gov [41] and a Data Safety Monitoring Board maintained oversight of its implementation.

## Results

There were 2365 subjects enrolled from 218 villages between January 1, 2014 and September 30, 2017, yielding 1232 subjects in COACH intervention villages and 1133 in eCAU. All village clinics remained actively engaged in the trial throughout 12 months of participation with 85 and 67 subjects lost to follow-up in the COACH and eCAU groups respectively. There were no reportable serious adverse events. Fig 1 depicts recruitment and retention over that time frame. Table 1 provides the demographic and baseline characteristics of the samples. Because of baseline differences between groups at the p<.05 level, religion, employment status, all WHOQOL-BREF scales, MOS-SSS-C, Social Network Size, count of comorbidities, and HTN control were included as covariates in subsequent analyses.

**Table 1.**
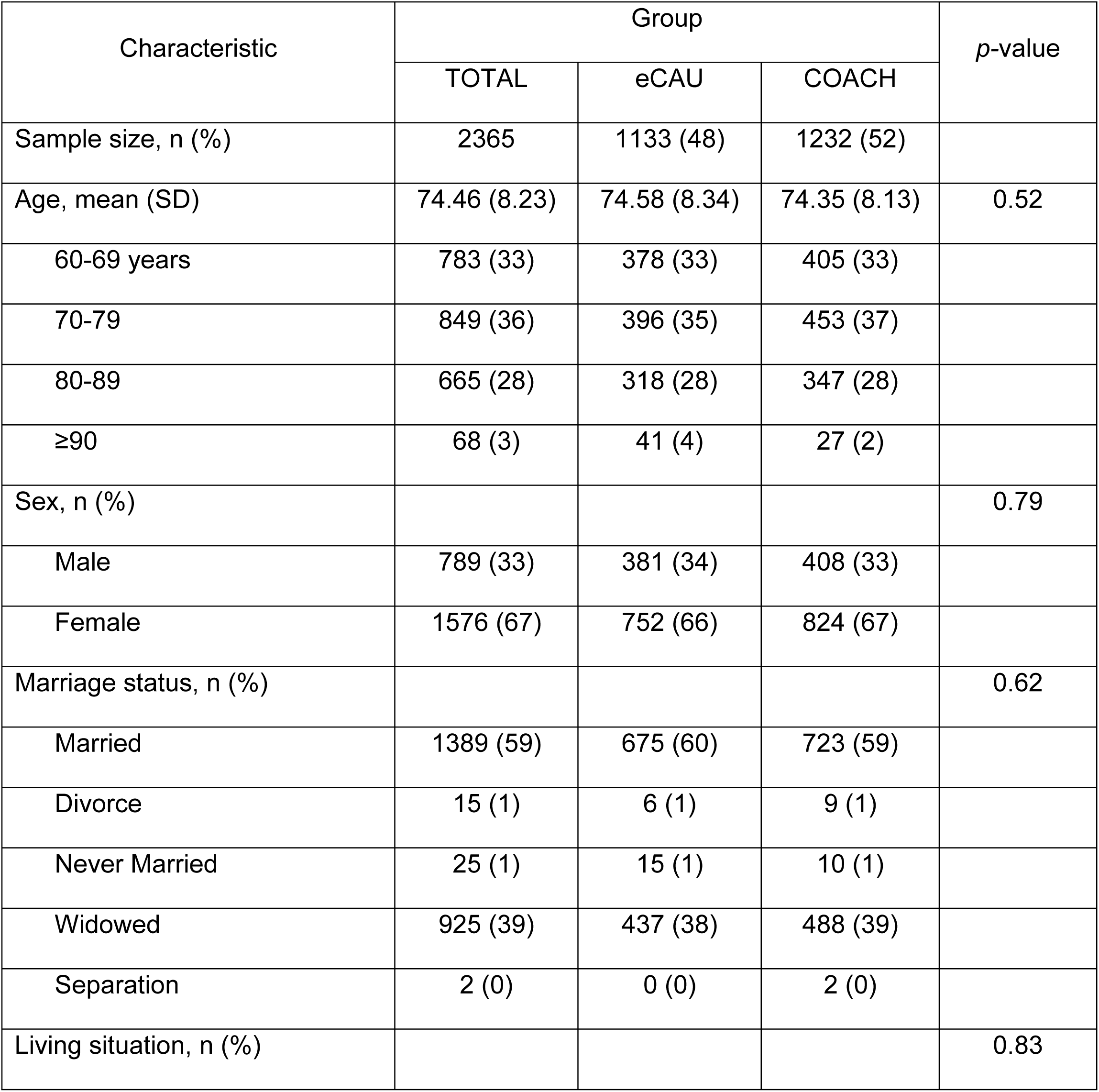
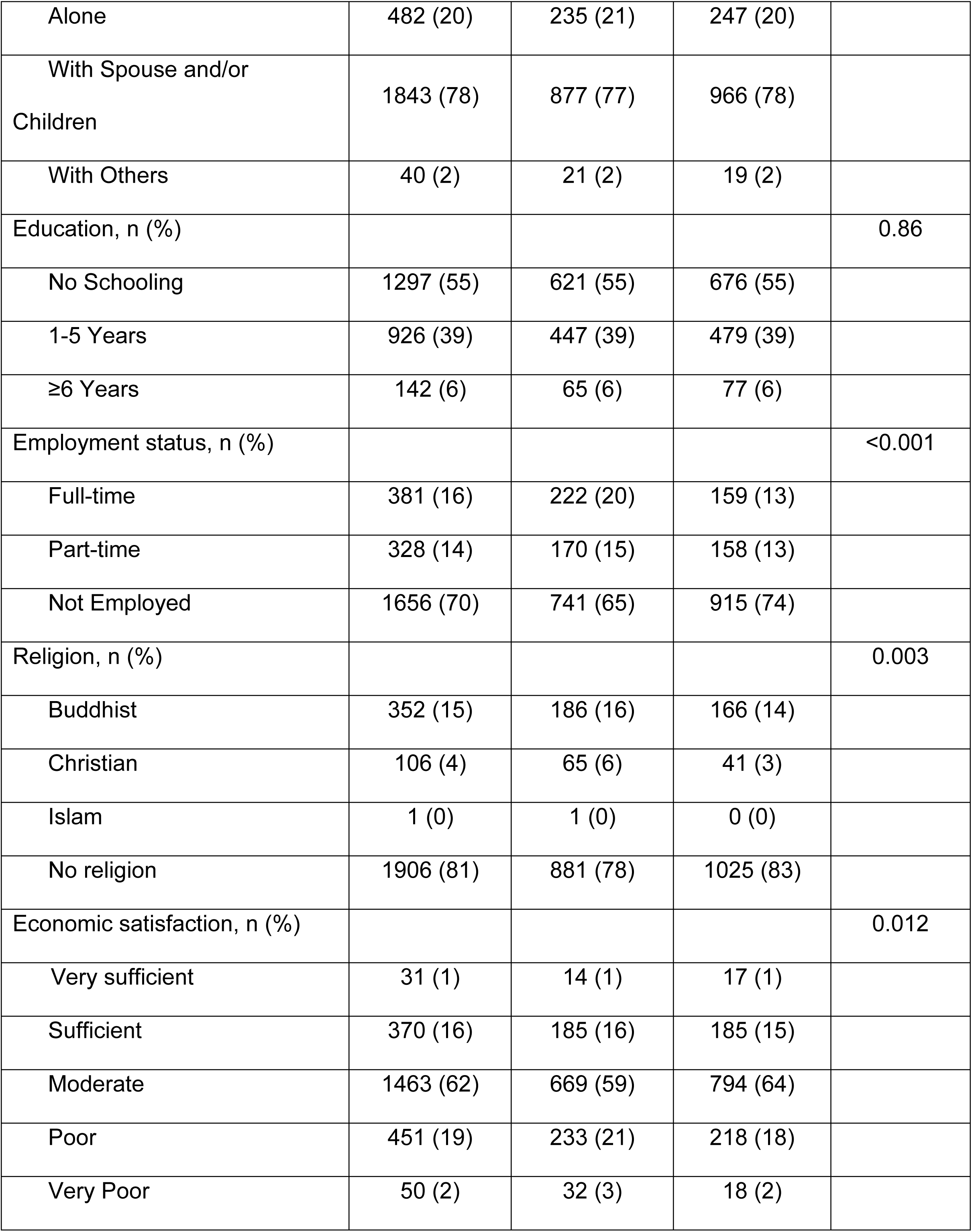
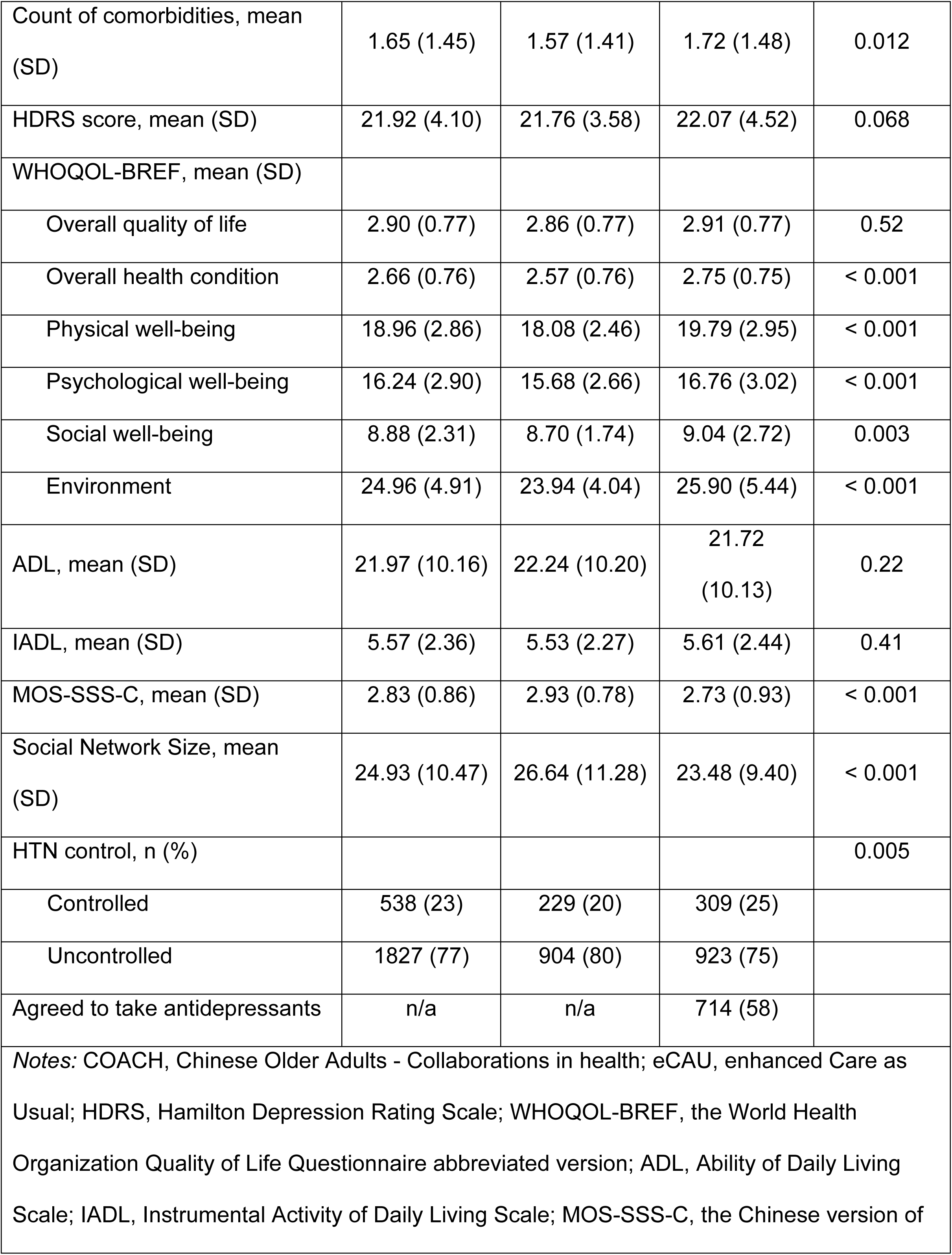
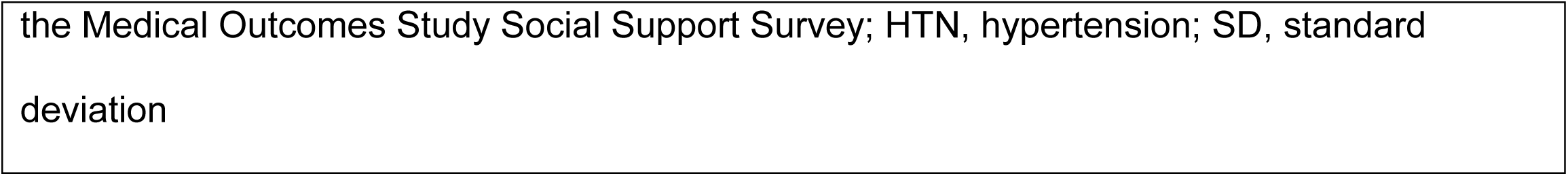
Demographic and clinical characteristics of subjects at baseline (N=2365)

| Characteristic | Group |  |  | p-value |
| --- | --- | --- | --- | --- |
|  | TOTAL | eCAU | COACH |  |
| Sample size, n (%) | 2365 | 1133 (48) | 1232 (52) |  |
| Age, mean (SD) | 74.46 (8.23) | 74.58 (8.34) | 74.35 (8.13) | 0.52 |
| 60-69 years | 783 (33) | 378 (33) | 405 (33) |  |
| 70-79 | 849 (36) | 396 (35) | 453 (37) |  |
| 80-89 | 665 (28) | 318 (28) | 347 (28) |  |
| ≥90 | 68 (3) | 41 (4) | 27 (2) |  |
| Sex, n (%) |  |  |  | 0.79 |
| Male | 789 (33) | 381 (34) | 408 (33) |  |
| Female | 1576 (67) | 752 (66) | 824 (67) |  |
| Marriage status, n (%) |  |  |  | 0.62 |
| Married | 1389 (59) | 675 (60) | 723 (59) |  |
| Divorce | 15 (1) | 6 (1) | 9 (1) |  |
| Never Married | 25 (1) | 15 (1) | 10 (1) |  |
| Widowed | 925 (39) | 437 (38) | 488 (39) |  |
| Separation | 2 (0) | 0 (0) | 2 (0) |  |
| Living situation, n (%) |  |  |  | 0.83 |
| Alone | 482 (20) | 235 (21) | 247 (20) |  |
| With Spouse and/or<br>Children | 1843 (78) | 877 (77) | 966 (78) |  |
| With Others | 40 (2) | 21 (2) | 19 (2) |  |
| Education, n (%) |  |  |  | 0.86 |
| No Schooling | 1297 (55) | 621 (55) | 676 (55) |  |
| 1-5 Years | 926 (39) | 447 (39) | 479 (39) |  |
| ≥6 Years | 142 (6) | 65 (6) | 77 (6) |  |
| Employment status, n (%) |  |  |  | <0.001 |
| Full-time | 381 (16) | 222 (20) | 159 (13) |  |
| Part-time | 328 (14) | 170 (15) | 158 (13) |  |
| Not Employed | 1656 (70) | 741 (65) | 915 (74) |  |
| Religion, n (%) |  |  |  | 0.003 |
| Buddhist | 352 (15) | 186 (16) | 166 (14) |  |
| Christian | 106 (4) | 65 (6) | 41 (3) |  |
| Islam | 1 (0) | 1 (0) | 0 (0) |  |
| No religion | 1906 (81) | 881 (78) | 1025 (83) |  |
| Economic satisfaction, n (%) |  |  |  | 0.012 |
| Very sufficient | 31 (1) | 14 (1) | 17 (1) |  |
| Sufficient | 370 (16) | 185 (16) | 185 (15) |  |
| Moderate | 1463 (62) | 669 (59) | 794 (64) |  |
| Poor | 451 (19) | 233 (21) | 218 (18) |  |
| Very Poor | 50 (2) | 32 (3) | 18 (2) |  |
| Count of comorbidities, mean (SD) | 1.65 (1.45) | 1.57 (1.41) | 1.72 (1.48) | 0.012 |
| HDRS score, mean (SD) | 21.92 (4.10) | 21.76 (3.58) | 22.07 (4.52) | 0.068 |
| WHOQOL-BREF, mean (SD) |  |  |  |  |
| Overall quality of life | 2.90 (0.77) | 2.86 (0.77) | 2.91 (0.77) | 0.52 |
| Overall health condition | 2.66 (0.76) | 2.57 (0.76) | 2.75 (0.75) | < 0.001 |
| Physical well-being | 18.96 (2.86) | 18.08 (2.46) | 19.79 (2.95) | < 0.001 |
| Psychological well-being | 16.24 (2.90) | 15.68 (2.66) | 16.76 (3.02) | < 0.001 |
| Social well-being | 8.88 (2.31) | 8.70 (1.74) | 9.04 (2.72) | 0.003 |
| Environment | 24.96 (4.91) | 23.94 (4.04) | 25.90 (5.44) | < 0.001 |
| ADL, mean (SD) | 21.97 (10.16) | 22.24 (10.20) | 21.72 (10.13) | 0.22 |
| IADL, mean (SD) | 5.57 (2.36) | 5.53 (2.27) | 5.61 (2.44) | 0.41 |
| MOS-SSS-C, mean (SD) | 2.83 (0.86) | 2.93 (0.78) | 2.73 (0.93) | < 0.001 |
| Social Network Size, mean (SD) | 24.93 (10.47) | 26.64 (11.28) | 23.48 (9.40) | < 0.001 |
| HTN control, n (%) |  |  |  | 0.005 |
| Controlled | 538 (23) | 229 (20) | 309 (25) |  |
| Uncontrolled | 1827 (77) | 904 (80) | 923 (75) |  |
| Agreed to take antidepressants | n/a | n/a | 714 (58) |  |

*Notes:* COACH, Chinese Older Adults - Collaborations in health; eCAU, enhanced Care as Usual; HDRS, Hamilton Depression Rating Scale; WHOQOL-BREF, the World Health Organization Quality of Life Questionnaire abbreviated version; ADL, Ability of Daily Living Scale; IADL, Instrumental Activity of Daily Living Scale; MOS-SSS-C, the Chinese version of
the Medical Outcomes Study Social Support Survey; HTN, hypertension; SD, standard deviation

**Fig 1.**
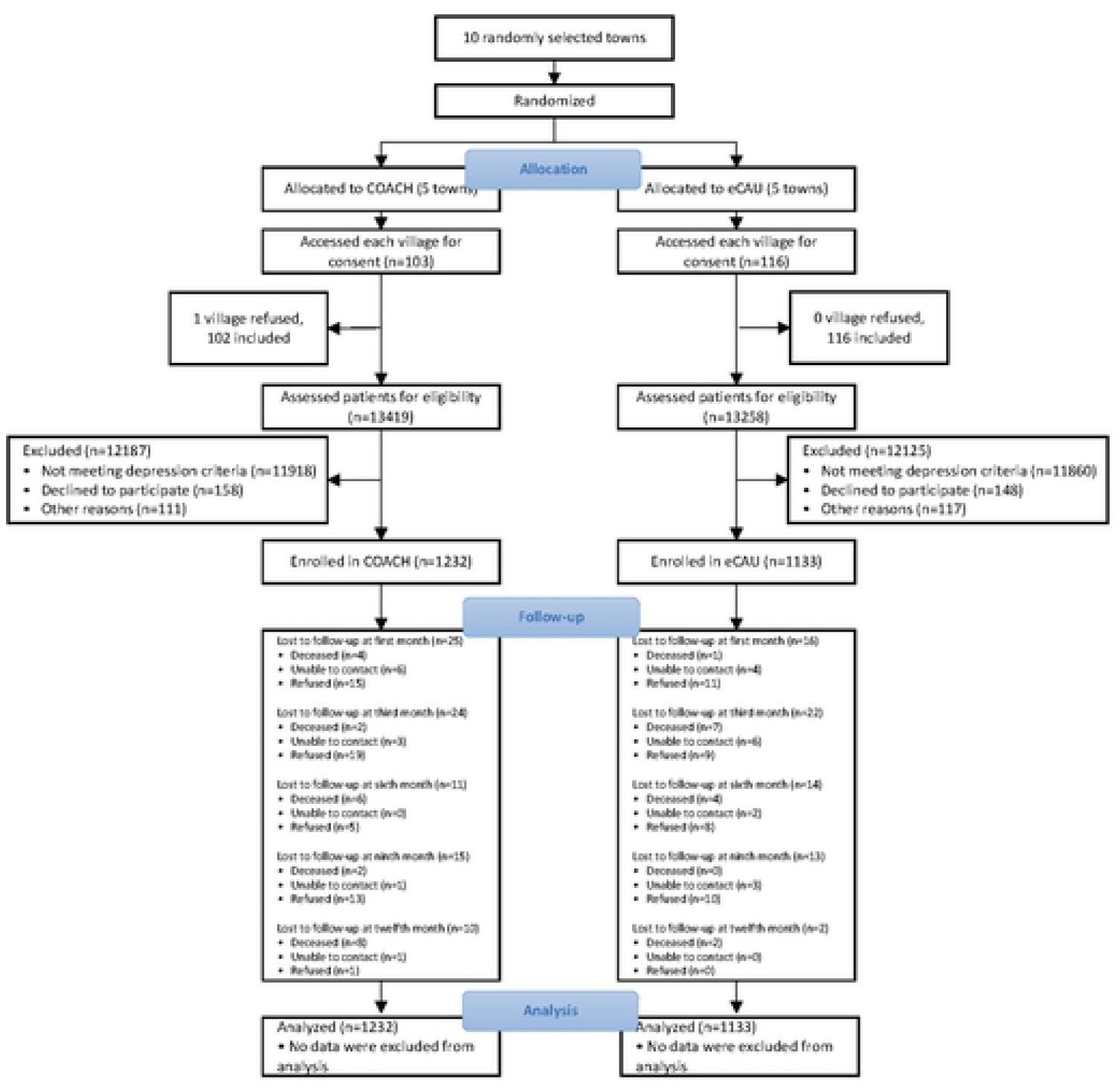
CONSORT diagram of COACH Study subject flow.

As depicted in Table 2, HDRS total scores (SD) decreased steadily over 12 months of study participation for those receiving COACH, albeit to levels still considered to be moderately symptomatic at 12.7 (4.2). eCAU subjects had a smaller reduction in HDRS score over 12 months from 21.8 (3.6) to 18.8 (4.7). The group x time interaction was significant with a large effect size, suggesting that the COACH group had a faster reduction in depressive symptom severity than the eCAU group. HTN control was more likely to be achieved by subjects in COACH villages, increasing from 25.1% to 71.6% over the 12 months of study participation, than those who received eCAU (from 20.2% to 40.9%). The group x time interaction was significant with a large effect size for HTN control as well.

**Table 2.**
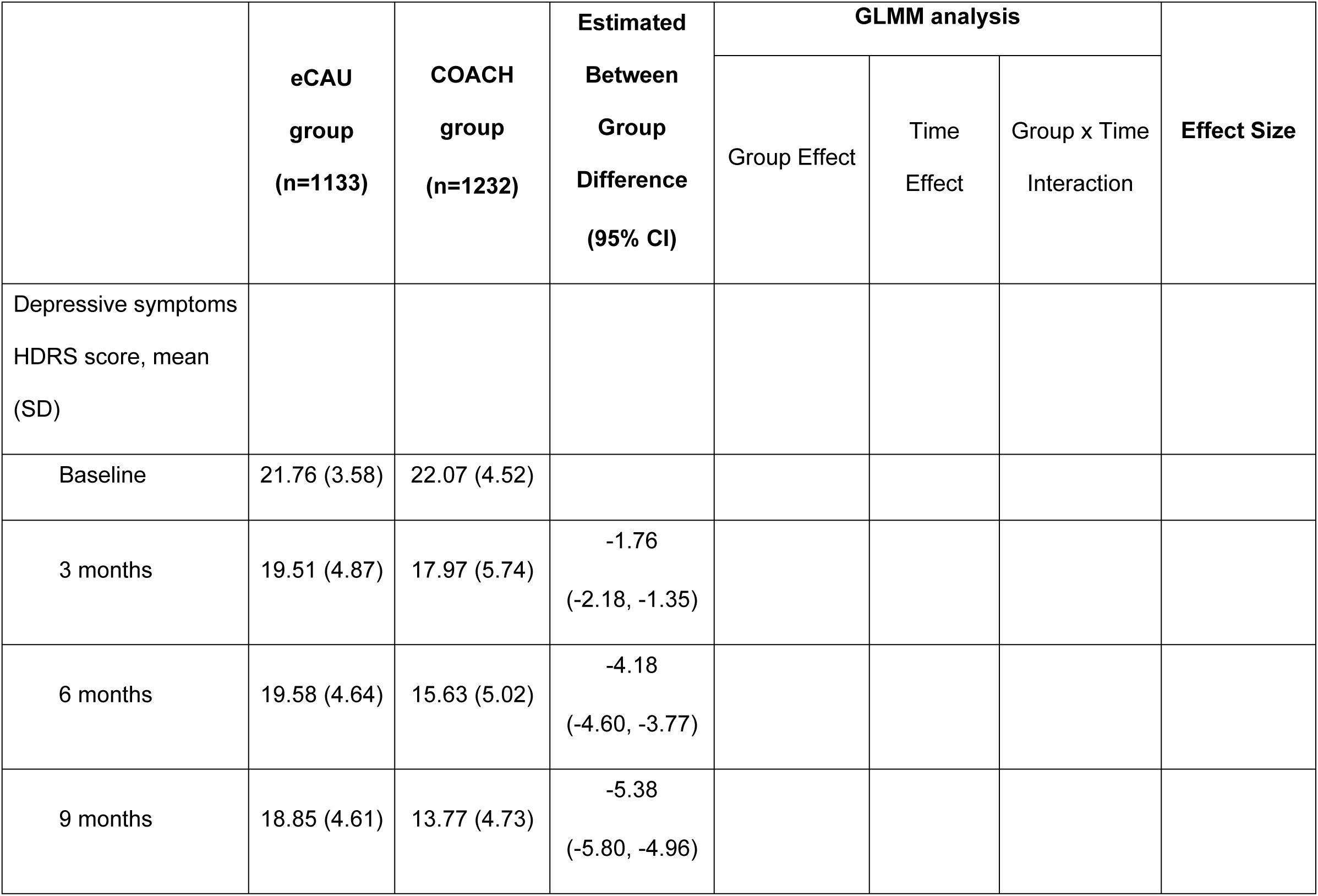
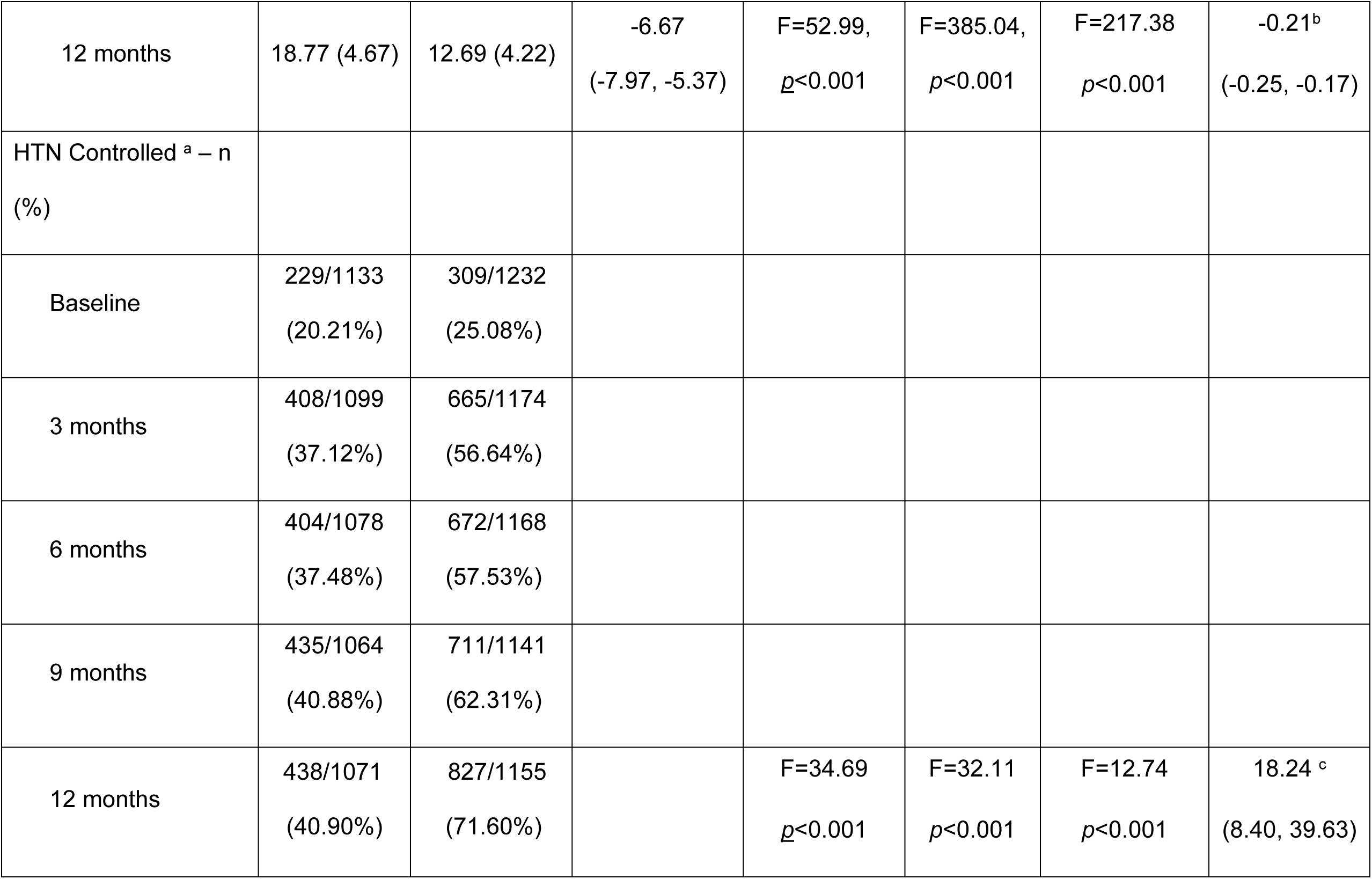
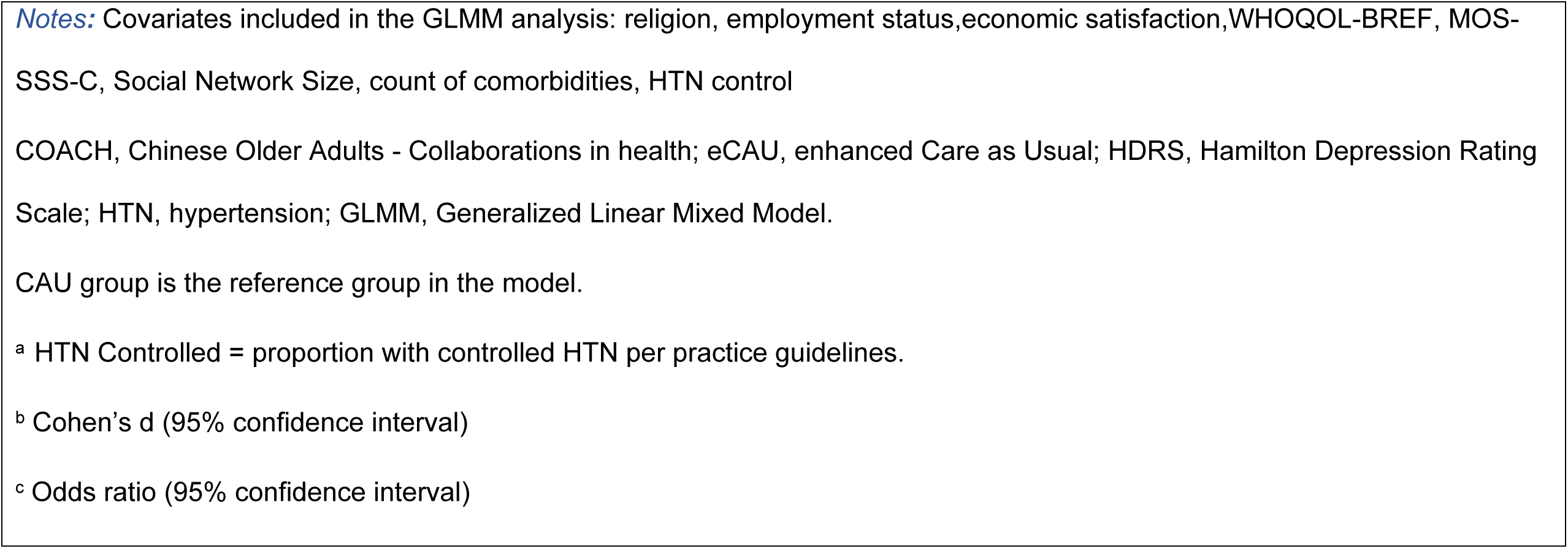
Depression and HTN outcomes in eCAU and COACH groups.

|  | **eCAU**<br>**group**<br>**(n=1133)** | **COACH**<br>**group**<br>**(n=1232)** | **Estimated**<br>**Between**<br>**Group**<br>**Difference**<br>**(95% CI)** | **GLMM analysis** |  |  | **Effect Size** |
| --- | --- | --- | --- | --- | --- | --- | --- |
|  |  |  |  | Group Effect | Time<br>Effect | Group x Time<br>Interaction |  |
| Depressive symptoms<br>HDRS score, mean<br>(SD) |  |  |  |  |  |  |  |
| Baseline | 21.76 (3.58) | 22.07 (4.52) |  |  |  |  |  |
| 3 months | 19.51 (4.87) | 17.97 (5.74) | -1.76<br>(-2.18, -1.35) |  |  |  |  |
| 6 months | 19.58 (4.64) | 15.63 (5.02) | -4.18<br>(-4.60, -3.77) |  |  |  |  |
| 9 months | 18.85 (4.61) | 13.77 (4.73) | -5.38<br>(-5.80, -4.96) |  |  |  |  |
| 12 months | 18.77 (4.67) | 12.69 (4.22) | -6.67<br>(-7.97, -5.37) | F=52.99,<br>$p<0.001$ | F=385.04,<br>$p<0.001$ | F=217.38<br>$p<0.001$ | -0.21<sup>b</sup><br>(-0.25, -0.17) |
| HTN Controlled <sup>a</sup> – n<br>(%) |  |  |  |  |  |  |  |
| Baseline | 229/1133<br>(20.21%) | 309/1232<br>(25.08%) |  |  |  |  |  |
| 3 months | 408/1099<br>(37.12%) | 665/1174<br>(56.64%) |  |  |  |  |  |
| 6 months | 404/1078<br>(37.48%) | 672/1168<br>(57.53%) |  |  |  |  |  |
| 9 months | 435/1064<br>(40.88%) | 711/1141<br>(62.31%) |  |  |  |  |  |
| 12 months | 438/1071<br>(40.90%) | 827/1155<br>(71.60%) | | F=34.69<br>$p<0.001$ | F=32.11<br>$p<0.001$ | F=12.74<br>$p<0.001$ | 18.24 <sup>c</sup><br>(8.40, 39.63) |

**Notes:** Covariates included in the GLMM analysis: religion, employment status, economic satisfaction, WHOQOL-BREF, MOS-SSS-C, Social Network Size, count of comorbidities, HTN control
COACH, Chinese Older Adults - Collaborations in health; eCAU, enhanced Care as Usual; HDRS, Hamilton Depression Rating Scale; HTN, hypertension; GLMM, Generalized Linear Mixed Model.
CAU group is the reference group in the model.
<sup>a</sup> HTN Controlled = proportion with controlled HTN per practice guidelines.
<sup>b</sup> Cohen's d (95% confidence interval)
<sup>c</sup> Odds ratio (95% confidence interval)

A substantial minority (n=518; 42%) of COACH group participants declined to take antidepressant medications as part of their treatment. Comparing those who did accept antidepressants (Antidep[+]) with those who did not (Antidep[-]), there were no notable differences in demographic and clinical characteristics at baseline (S1 Table). Figs 2 and 3 allow for visual comparison of eCAU with Antidep[+] and Antidep[-] subgroups. The COACH Antidep[-] group had a pattern of response in HDRS total score between that of eCAU and Antidep[+] (Fig 2). With regard to HTN control, a different pattern emerged wherein improvements in HTN control were clearly greater for both COACH subgroups than for eCAU subjects (Fig 3). The strong impact of the intervention appears equivalent with regard to HTN for both COACH subgroups relative to eCAU.

**Fig 2.**
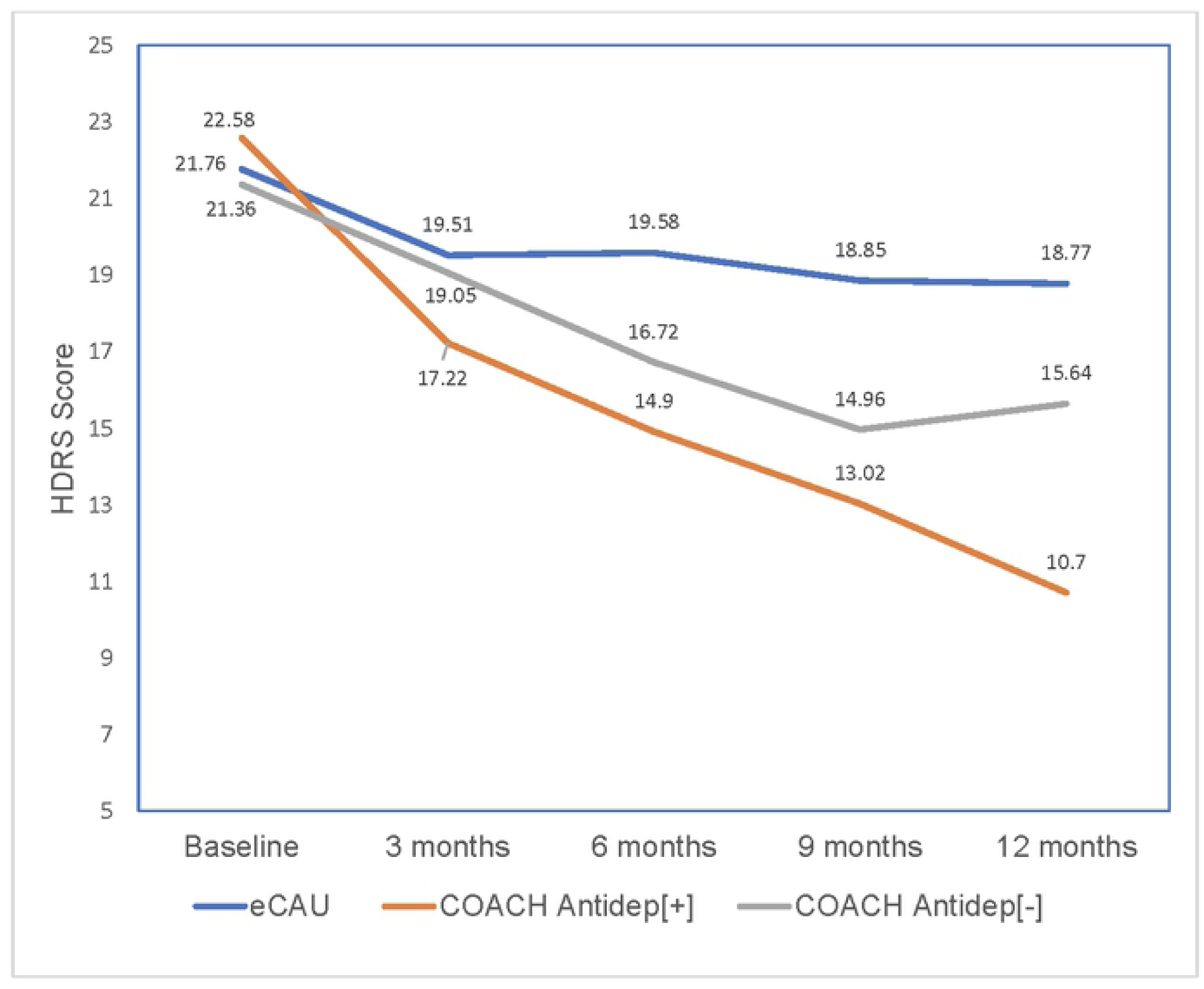
Depressive symptom severity over 12 months of 3 groups of participants. eCAU, COACH subjects who accepted antidepressant medications (Antidep[+]); COACH subjects who declined antidepressant medications (Antidep[-]). HDRS = Hamilton Depression Rating Scale

**Fig 3.**
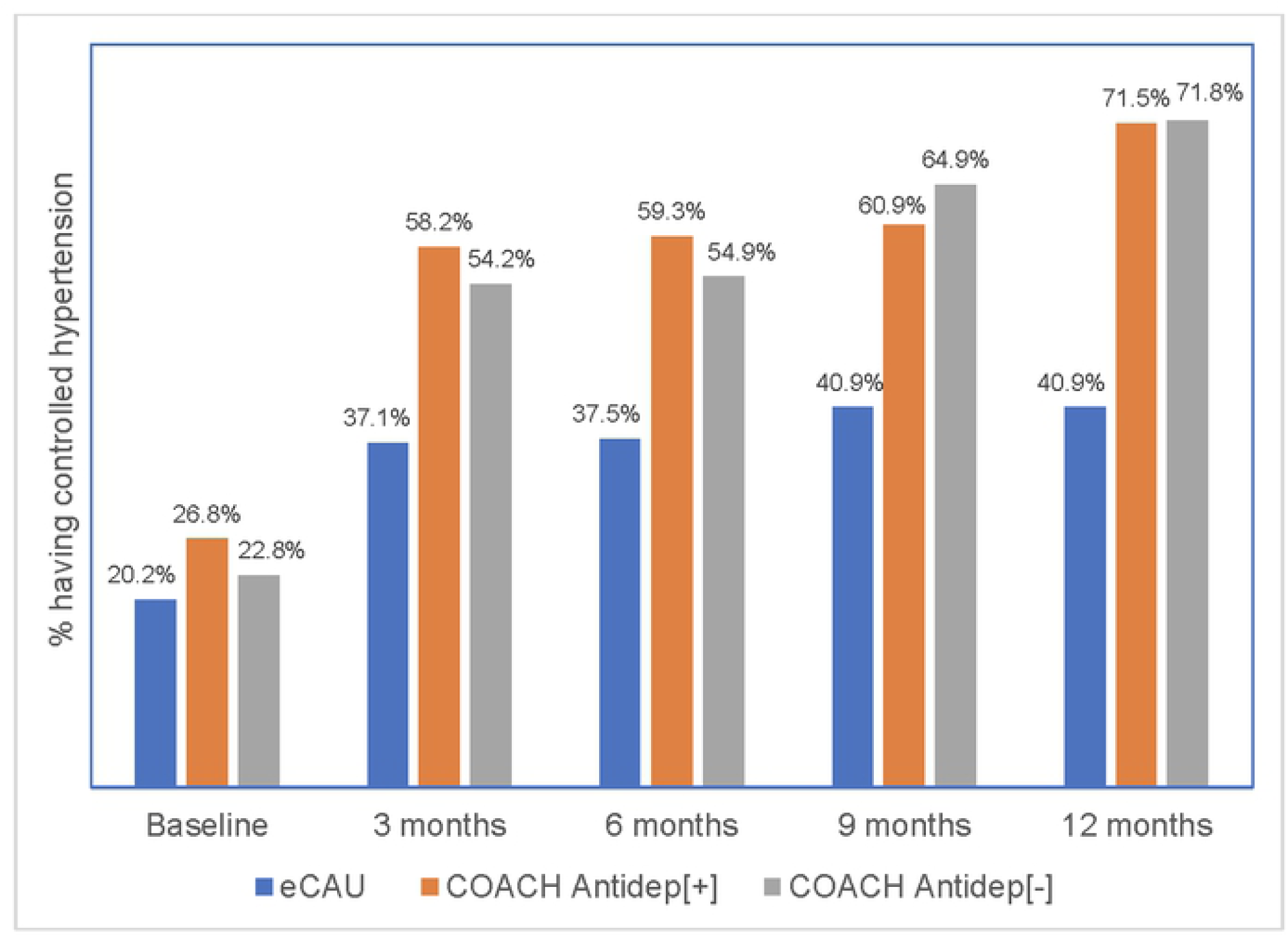
Proportion of subjects in 3 groups who achieved hypertension (HTN) control over 12 months. eCAU; COACH subjects who accepted antidepressant medications (Antidep[+]); COACH subjects who declined antidepressant medications (Antidep[-])

## Discussion

Studies in the West indicate that comorbidity of HTN and depression compounds their adverse impacts, and that integrated depression care management in primary care settings improves outcomes of both conditions. We are aware of only two randomized controlled trials that have examined collaborative care management of comorbid medical and mental disorders in developing countries. One conducted in urban Indian primary care clinics found greater reduction in depressive symptoms and improved type 2 diabetes control among adults who received a multi-component collaborative care intervention than among those who received care as usual [42]. The second employed a collaborative care model for management of comorbid depression and HTN in South African primary care clinics, finding no benefit compared to CAU [43]. Neither study examined older adults.

Consistent with our hypotheses, we found that older rural village clinic patients who received the COACH intervention had significantly greater improvement in depressive symptoms and HTN control than did those in village clinics providing eCAU. The integrated care model shown to be effective in Western primary care settings appears to have promise in rural China for depressed older adults with comorbid medical illness.

Although these findings were highly significant with large effect sizes, the level of improvement in depression outcomes was lower among COACH subjects than had been observed in our earlier study in urban primary care clinics, even though collaborative care management with the same antidepressant medication algorithm was used in both [22]. In rural clinics delivering the COACH intervention, subjects had on average a 43% reduction in HDRS score, whereas older adults who received depression care management using the same antidepressant medication regimen in urban clinics achieved a mean 66% reduction of HDRS total scores over 12 months. Post hoc reasoning suggested that the difference could be accounted for by more uniform antidepressant use in the urban clinic subjects (100% received antidepressant treatment vs. 58% in COACH). Because the study design did not randomize antidepressant exposure, we could not directly test this hypothesis in the COACH study. However, comparison of the eCAU, Antidep[-], and Antidep[+] groups is suggestive (Fig 2), showing proportional reductions in average HDRS score over 12 months of 14%, 28%, and 53% in the three groups respectively. Further study is needed to establish the priority that should be placed on provision of antidepressants to older adults with clinically significant depressive symptoms in under-resourced areas, the great majority of whom will have had no prior exposure to medication treatment.

Limitations of the study include generalizability of the model to other regions of China or other LMICs [44]. As well, there were methodological weaknesses to which RCTs in rural, under-resourced areas are vulnerable. We were unable to mask research assessors to which condition a village was assigned. Although they were kept unaware of the study’s hypotheses, there is the risk of assessment bias. Because the study randomized five towns to each intervention group while power calculations were made using village as the unit of randomization, the trial could have been underpowered depending on the magnitude of between cluster differences. As anticipated based on the study data, however, the ICC for the HDRS at the village level (0.064) was greater than at the town level ICC (0.027), well inside our assumed range for the power calculation. Also, there was no meaningful difference between the ICCs for HTN control at the town and village levels (0.025 and 0.021 respectively) and the observed effect sizes were large, indicating that the study is sufficiently powered.

We do not know the extent to which subjects followed lifestyle recommendations for management of HTN and depression such as diet, exercise, and socialization. We were unable to assess adherence to medications as planned in the original proposal, precluding examination of the temporal relationships between intervention exposure and changes in depression and BP control in COACH and eCAU groups.

While the COACH intervention appears effective as administered, study is needed of modifications that may further increase its impact, such as employment of telehealth technology to link rural village-based COACH teams to psychiatric supports remotely [45], and training of AWs in evidence-based psychotherapies and behavior change techniques to promote a healthy lifestyle [46].

## Data Availability

We are currently preparing to submit the data to Harvard Dataverse. The accession numbers or DOI will be assigned at a later date when we complete the process.

## Acknowledgments

We thank the rural village residents of Tonglu and Jiande Counties who participated in the study, their primary care doctors, and aging workers for their contributions.

